# Tracking the emergence of disparities in the subnational spread of COVID-19 in Brazil using an online application for real-time data visualisation: a longitudinal analysis

**DOI:** 10.1101/2021.04.30.21256386

**Authors:** Paul Mee, Neal Alexander, Philippe Mayaud, Felipe de Jesus Colon Gonzalez, Sam Abbott, Andreza Aruska de Souza Santos, Andre Luis Acosta, Kris V. Parag, Rafael H. M. Pereira, Carlos A. Prete, Ester C. Sabino, Nuno R. Faria, LSHTM Centre for Mathematical Modelling of Infectious Disease COVID-19 working group, Oliver J Brady

## Abstract

**Background:** Brazil is one of the countries worst affected by the COVID-19 pandemic with over 20 million cases and 557,000 deaths reported. Comparison of real-time local COVID-19 data between areas is essential for understanding transmission, measuring the effects of interventions and predicting the course of the epidemic, but are often challenging due to different population sizes and structures.

**Methods:** We describe the development of a new app for the real-time visualisation of COVID-19 data in Brazil at the municipality level. In the CLIC-Brazil app, daily updates of case and death data are downloaded, age standardised and used to estimate reproduction number (*R*_*t*_). We show how such platforms can perform real-time regression analyses to identify factors associated with the rate of initial spread and early reproduction number. We also use survival methods to predict the likelihood of occurrence of a new peak of COVID-19 incidence.

**Findings:** After an initial introduction in São Paulo and Rio de Janeiro states in early March 2020, the epidemic spread to Northern states and then to highly populated coastal regions and the Central-West. Municipalities with higher metrics of social development experienced earlier arrival of COVID-19 (decrease of 11·1 days [95% CI:13·2,8·9] in the time to arrival for each 10% increase in the social development index). Differences in the initial epidemic intensity (mean *Rt*) were largely driven by geographic location and the date of local onset.

**Interpretation:** This study demonstrates that platforms that monitor, standardise and analyse the epidemiological data at a local level can give useful real-time insights into outbreak dynamics that can be used to better adapt responses to the current and future pandemics.

**Funding:** This project was supported by a Medical Research Council UK (MRC-UK) -São Paulo Research Foundation (FAPESP) CADDE partnership award (MR/S0195/1 and FAPESP 18/14389-0)

## Introduction

COVID-19 is a new respiratory and multi-organ illness caused by infection with the severe acute respiratory syndrome coronavirus type-2 (SARS-CoV-2) which emerged in December 2019 in Wuhan, China. As of 4^th^ August 2021 over 200 million COVID-19 cases and over 4·2 million deaths have been reported worldwide ^1^. Brazil is one of the worst affected countries with over 20 million cases and 557,000 deaths reported ^1^. Heterogenous patterns of propagation of the virus across the country have been driven by a complex intersection of causative factors including; continued movements of people between urban centres throughout the epidemic, differential imposition of interventions designed to reduce transmission and relative isolation of municipalities from the major population centres^2^. Currently the country is experiencing a second wave of the epidemic driven by a viral variant which arose in the Amazonas region and has quickly spread throughout the country^3^.

Comparisons of incidence between different local areas can give important insights into the patterns of spread and the burden of an epidemic and help to separate generalisable from context specific transmission trends. Such comparisons are complicated by differences in the characteristics of local populations that affect the risk of disease even if levels of infection are equal. In particular, age is a major risk factor for infection with SARS-CoV-2 and subsequent COVID-19 disease. Consequently, differences in the age distribution need to be taken into account when comparing local areas,^4–6^ The differences in epidemic severity between places could also be driven by sociodemographic factors, ethnicity, the relative isolation of different regions and the levels of implementation and effectiveness of non-pharmaceutical interventions. A serosurvey conducted in May and June of 2020 in cities across Brazil found evidence that prevalence of SARS-Cov-2 antibodies, an indicator of prior infection, was higher for those; living in crowded conditions, of non-white ethnicity and those in the lowest socio-economic groups^7^. In contrast a study of data from the early stages of the epidemic in Brazil, up to May 2020, found that the those in higher socio-economic groups were more likely to have a positive test for COVID-19^8^. This may imply a changing risk profile over time or may reflect differential access to testing among socio-economic groups.

With the increased roll-out of vaccinations, local differences in vaccine uptake also need to be considered^9^. All of these aspects could impact on the rate of spread and onset time of the epidemic in a given locality. Quantifying the role of these components, and the interplay between them, is important for understanding patterns of past infection and the likely severity of future waves of infection.

The field of real-time analysis of infectious disease data is rapidly expanding, in part due to greater automation, digitisation and online sharing of data^10^. Projects such as the Johns Hopkins University Covid-19 Dashboard^11^. aim to provide a global overview of cases and deaths with the goal of making international comparisons^12^. and a number of sub-national-level real-time data dashboards have also been established for finer scale domestic comparisons such as that for Italy^13^. Such dashboards are useful for rapid situation reports, yet direct comparison between regions with differing age distributions and onset times offers limited epidemiological insight into the rate of spread and local burden of the epidemic.

Websites such as EpiForecasts^14,15^ the CDC Covid-19 Forecasting Hub^16–18^ and ‘Short-term forecasts for multiple countries’^19^ aim to make and compare short-term projections of disease incidence using mathematical and statistical models. As part of this process, some models aim to estimate the effective reproduction number *R*_*t*_, an estimate of the average number of new infections that will occur from each infected person. Real-time estimates of *R*_*t*_ over time^20,21^ are useful for planning interventions to mitigate the impact of the epidemic^22^. Accurate estimation of *R*_*t*_ is complicated by the need to correct for the delays between infection and reporting of cases of disease and under-ascertainment of cases. Due to the computational resources required to run these forecasting models, most existing analysis dashboards only give predictions at the national or first administrative level (e.g. State in Brazil) and are not updated daily to reflect the latest situation^23^.

There is a need for a new class of dashboards that are able to perform basic data standardisation to account for differing population age structures and allow for regional comparisons. Additionally, such dashboards should provide local summaries of key epidemiological parameters and support rapid data analyses of outbreak dynamics whilst retaining the contemporary focus of real-time data streams. In response to this we have developed an online application for the real-time visualisation of COVID-19 cases and death data in Brazil at the municipality (second-level administrative division) level. This allows real-time comparisons of the development of the epidemic in Brazil to be made at a local level to allow local decision makers to track and compare epidemic progression rates between different areas. The COVID-19 Local Information Comparison (CLIC Brazil) [https://cmmid.github.io/visualisations/lacpt] has been active since May 2020, early in the Brazilian epidemic. The data underlying the app are updated daily and relevant local data summaries and analytics re-computed. Here we describe the CLIC Brazil application and the insights about the early evolution of the COVID-19 outbreak in Brazil that it has helped reveal.

## Methods

### Context

Brazil is the largest and most populous country in South and Latin America, with a total population estimated at over 213 million in 2021^24^. The country is composed of 26 states and the Federal District and 5570 municipalities.

### Data Sources

The numbers of COVID-19 cases and deaths, aggregated by municipality were automatically downloaded daily from the Brazil.io COVID-19 project repository^25^. This repository contains data extracted from the bulletins of state health secretariats. Data on the distribution of the population by age and the sociodemographic characteristics of each municipality were obtained from the most recent national demographic census, run by the Instituto Brasileiro de Geografia e Estatística (IBGE) in 2010^26^. To allow age standardisation of cases, data on the age distribution of COVID-19 cases were derived from case reports throughout Brazil between 2^nd^ February and 25^th^ March 2020, collected by the Brazilian Ministry of Health and were used with their permission. Using this data enabled consistent standardisation throughout the epidemic. In order to assess the effect of geographical remoteness from major cities, the travel time in hours from each municipality to the most populous metropolitan area in the state was calculated using WorldPop population data^27,28^ and the Malaria Atlas Project travel time friction surface using the Malaria Atlas^28^ accumulated cost route finding algorithm within the MalariaAtlas R package^29,30^. The socio-demographic Index (SDI) is a composite average of the rankings of the incomes per capita, average educational attainment, and fertility rates scaled between 0 (lowest) and 1 (highest)^31^. The geographic region in which each place was located was assigned using a standardised designation which groups the States and the Federal District of Brasilia into five macro regions of national planning^32^ (Fig 1). Data uploaded to Brazil.IO between 25-Feb-2020 and 14-Jul-2021 was used for the analyses reported here, updated data can be downloaded from the CLIC-Brazil app. Data on the types of non-pharmaceutical interventions implemented, and the dates of their announcement were extracted from data collated by the Cepal Observatory with edits and updates on timing of interventions at the municipality and state level by de Souza Santos et al ^3,8^. These were used to compare the dates for the implementation and arrival of the epidemic locally. Full details of the data sources and data processing are included in the Supplementary Materials.

**Figure 1:**
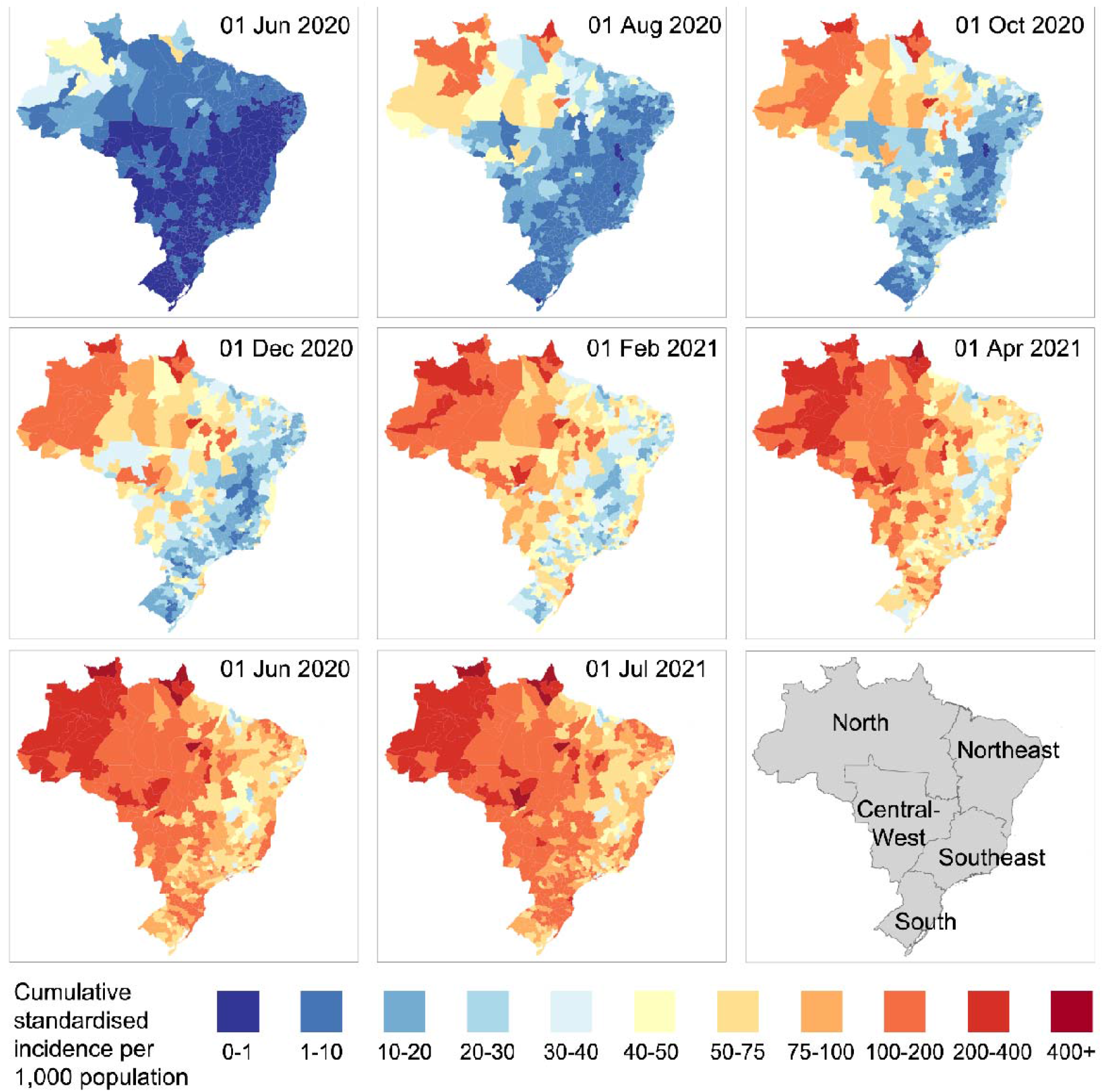
Spatial progression of the COVID-19 epidemic in Brazil. Standardised case incidence is a measure that allows comparison between municipalities with different population sizes and age structures. Standardised incidence for all 5570 municipalities has been aggregated to microregion (n = 557) by population-weighted averaging for visualisation purposes. The bottom right panel includes a map of the six macro regions of Brazil for reference.

### Features of the application

The homepage of the app shows a map of Brazil showing the number of cases in each municipality and a timeline for the development of the national epidemic. A series of tabs provide the following functionality to users: a comparison of standardised COVID-19 incidence between municipalities, trends in the association between sociodemographic variables and incidence, changes in *R*_*t*_ between selected municipalities over time, predictions of the likelihood that a particular municipality has reached peak incidence and the ability to download incidence estimates, *R*_*t*_ predictions and sociodemographic variables.

### Development of the application

The COVID-19 Local Information Comparison (CLIC Brazil) application was developed using the R package “shiny”, version 1.5.0 ^33^. CLIC Brazil provides users with options for graphical display of information and all computation required is handled remotely. Plots are generated using the R package “ggplot2” version 3.3.2 ^34^. Spatial data presented in the form of maps are portrayed using the “leaflet” R package version 2.0.3 ^35^. Screenshots from the app are shown in Figures S3 to S5 in the Supplementary materials. All code described in the paper can be downloaded from this github repository - https://github.com/Paul-Mee/clic_brazil.

### Analytical methods

#### Calculating the comparable measure of standardised incidence

To enable comparison of COVID-19 case counts between municipalities with different age structures and thus different probabilities of disease given infection, we standardised each municipality’s incidence to the national-level age structure. (See Supplementary Materials for details). When comparing the progression of the epidemic in different municipalities over time, a comparable outbreak start criterion has to be established. We defined the arrival of COVID-19 in a given municipality as the day in which cumulative standardised incidence first exceeded 1 case per 10,000 residents.

#### *R*_*t*_ Estimation

The raw case count data was adjusted to account for the differential probabilities of COVID-19 cases being reported on each day of the week (heaping). (See Supplementary Materials for details). The EpiFilter algorithm was used to estimate *R*_*t*_ ^36^, this method uses a recursive Bayesian filter to derive estimates from a time series of all incident cases. The serial interval (SI), defined as the time between the onset of symptoms in the source of infection and in the recipient, was modelled as a gamma distribution with a mean of 6·5 days with a standard deviation of 4·03 days^37^. A fixed value of 10 days was used for the delay between symptom onset and case reporting, this was consistent with an average value of 10·2 days for 2,420,904 suspected COVID-19 cases reported between March 1^st^ and August 18^th^ 2020 in all the state capitals and Federal District of Brazil in the e-SUS notification system^38^. Predictions were only made for municipalities with more than 30 days of data and more than 200 COVID-19 cases reported, to allow sufficient data for the algorithm to give reliable estimates^39^. The resultant Rt curves were plotted to enable comparison between selected cities in the CLIC Brazil app.

#### Regression analyses

Using data from the CLIC Brazil app, two regression models were formulated to quantify how the timing of arrival and growth rate of the COVID-19 epidemic in each municipality could be explained by sociodemographic characteristics and spatial connectivity. Initially a series of univariable regression models were developed to test whether each covariate was individually associated with the outcome. The variables included were; population density, the percentage of residences with i) piped water and ii) piped sewage, the travel time to the largest city in the state and the socio-demographic index (See Data Sources section in the Methods). Additionally the geographic region (Central-West, North, North-East, South, South-East) in which the municipality was located was included as a fixed effect to partially for residual confounding based on other unmeasured geographically associated characteristics. Following this, both forward and backward stepwise regression approaches were used to develop multivariable models. (See Supplementary Materials.)

#### Analysis of factors associated with time for the epidemic to arrive in a municipality

To assess which characteristics of a municipality were associated with arrival time of COVID-19 we first defined date of arrival in each municipality as the date when standardised incidence first exceeded 1 case per 10,000 residents. These dates where then compared to the date of arrival of COVID-19 in Brazil which we define as 31^st^ March 2020 (the date on which the first municipality exceeded an incidence of 1 per 10,000 residents) to calculate the number of days for arrival which formed the response variable for the Tobit regression analysis^40^ as implemented in the R package VGAM^41^. A Tobit regression formulation was necessary due to censoring, i.e. unknown days for arrival for municipalities that were not yet infected. To assess the sensitivity of our findings to our definition of COVID-19 arrival we repeated our analysis with a range of threshold incidence values (5 to 15 cases per 10,000).

#### Analysis of factors associated with the rate of growth in the early stages of the epidemic

To assess the which factors were associated with growth rate of epidemic in municipalities after COVID-19 had arrived, we calculated the mean value of *R*_*t*_ over the period 30-150 days post arrival in each municipality. Calculating *R*_*t*_ using data within this time window balanced the need to include enough data for accurate estimation^39^ with the need to estimate *R*_*t*_ before the build-up of substantial immunity or reactive interventions (therefore approximating *R*_*0*_). To test the sensitivity of our findings to the chosen width of this estimation window, we repeat the analysis with the end point for the mean *R*_*t*_ estimation varying between (100 and 180 days). Estimated mean *R*_*t*_ was then included as the response variable in a standard log-linear regression model using the “glm” function in base R with covariate selection as described above. In addition to the set of covariates described above we included the calendar time period in which the local epidemic commenced to control for residual temporal confounding. Initial univariate analyses suggested grouping calendar time period into three roughly equal categories, all within 2020 (14^th^ March to 1^st^ May, 2^nd^ May to 21^st^ May and 22^nd^ May to 6^th^ Nov) captured variation appropriately and that an interaction between calendar time period and geographic region should be considered as a separate (selectable) covariate.

#### Predicting whether a new maximum incidence will occur

Here we used Cox regression as implemented in the “coxph” function of the “survival” package in R ^42^ Click or tap here to enter text.to estimate the probability of each municipality surpassing its previous maximum weekly standardized incidence (i.e. a new “record” incidence) within the following 4 weeks. The analysis time was the number of weeks since the start of the epidemic (cumulative standardised incidence exceeded 1 case per 10,000 residents). The event of interest was the setting of a new record incidence, which in general occurs more than once. (See Supplementary Materials.)

The reporting guidelines in the reporting of studies Conducted using Observational Routinely-collected health Data (RECORD) Statement were used^43^. Click or tap here to enter text.The completed RECORD checklist is included in the Supplementary Materials.

## Results

To compare the spatial progression of the COVID-19 epidemic in Brazil, we mapped cumulative standardised disease incidence in Fig. 1

Despite the first introductions and local SARS-CoV-2 transmission events occurring in São Paulo and Rio de Janeiro states in early March 2020^44^ Click or tap here to enter text.the focus of the outbreak quickly shifted to the North region of the country where the first big outbreaks occurred in late May/early June, particularly in the border states of Amazonas, Roraima and Amapá (Fig. 1). By August 2020, COVID-19 transmission was widespread across the North region and began to spread to major coastal cities, particularly in the Northeast. By October, transmission had spread along the highly populated coastal areas and into the Central West region. Between August and October,SARS-CoV-2 spread to the final transmission free areas in sparsely populated inland regions and in the far South. By December 1^st^, transmission was widespread throughout the whole country. During November-December 2020 the re-emergence of large outbreaks occurred throughout the Central-West region alongside renewed growth in coastal cities of the South East and South. From February to July 2021, incidence remained high in the North and increased elsewhere, such that as of July 2021, most areas had cumulative standardised incidence rates comparable to some of the worst affected areas in the North.

To compare the trajectory of COVID-19 outbreaks in local areas once the first wave of epidemic had begun, we plotted cumulative standardised case counts per municipality in different states and regions in Brazil (Fig. 2). This revealed that the outbreak was comparatively faster growing and reached higher cumulative incidence in the North region of Brazil (Fig. 2A and F). Within this region, the most northerly states of Amazonas, Amapá and Roraima were the most severely affected with some municipalities experiencing cumulative case incidence rates as high as 45%. Areas in the Southeast and, until recently, South regions had slower growing outbreaks in the earlier stages of epidemic (Fig. 2D-F). Outbreak trajectories in the Central-West and Northeast regions were between the high rates observed in the North and low rates observed in the South in the first wave, but have since increased to the high levels seen elsewhere. (Fig. 2F). Despite these trends, there was considerable within region and within state heterogeneity in outbreak trajectories, suggesting factors other than just geographical location were important in shaping the trajectory of the epidemic. Plots for specific municipalities can be viewed in the CLIC Brazil app [https://cmmid.github.io/visualisations/lacpt]

**Figure 2:**
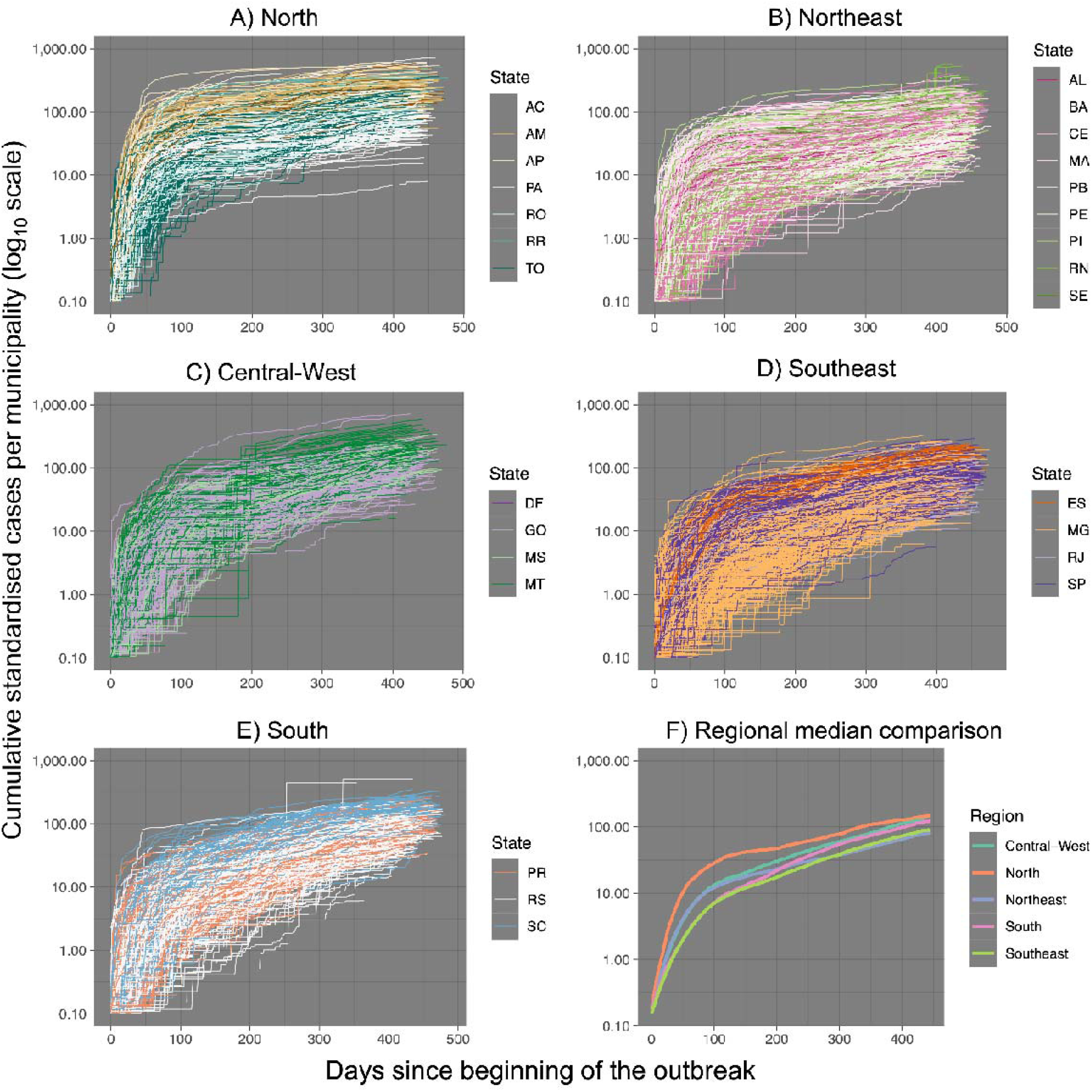
Comparison of outbreak trajectories in different regions of Brazil. Each line represents a municipality within the geographical area. Cases per municipality have been age- and population-standardised, plotted cumulatively and aligned to the date of detection of the first COVID-19 cases in each municipality (defined as an incidence of greater than 0·1 standardised cases per 1,000 residents). Only municipalities that have reported 50 or more cases are shown. Panel F compares region median trajectories.

Between 28^th^ February and 27^th^ March 2020, a range of state-level restrictions were announced to limit the spread of SARS-CoV2 including declaring an emergency, industry, retail, service and transport restrictions and school closures. At the time these interventions were announced, only a very small number of municipalities had reported a single COVID-19 case (51 of 5,570 municipalities). This indicates that the announcement of interventions in Brazil occurred before the first COVID-19 cases appeared in the majority of municipalities (mean of 54.8 days before the first case was reported in pre-emptive municipalities). Even in municipalities where interventions were announced reactively there was only a mean 2·2 days between reporting the first case and their announcement (red area in Fig. 3).

**Figure 3:**
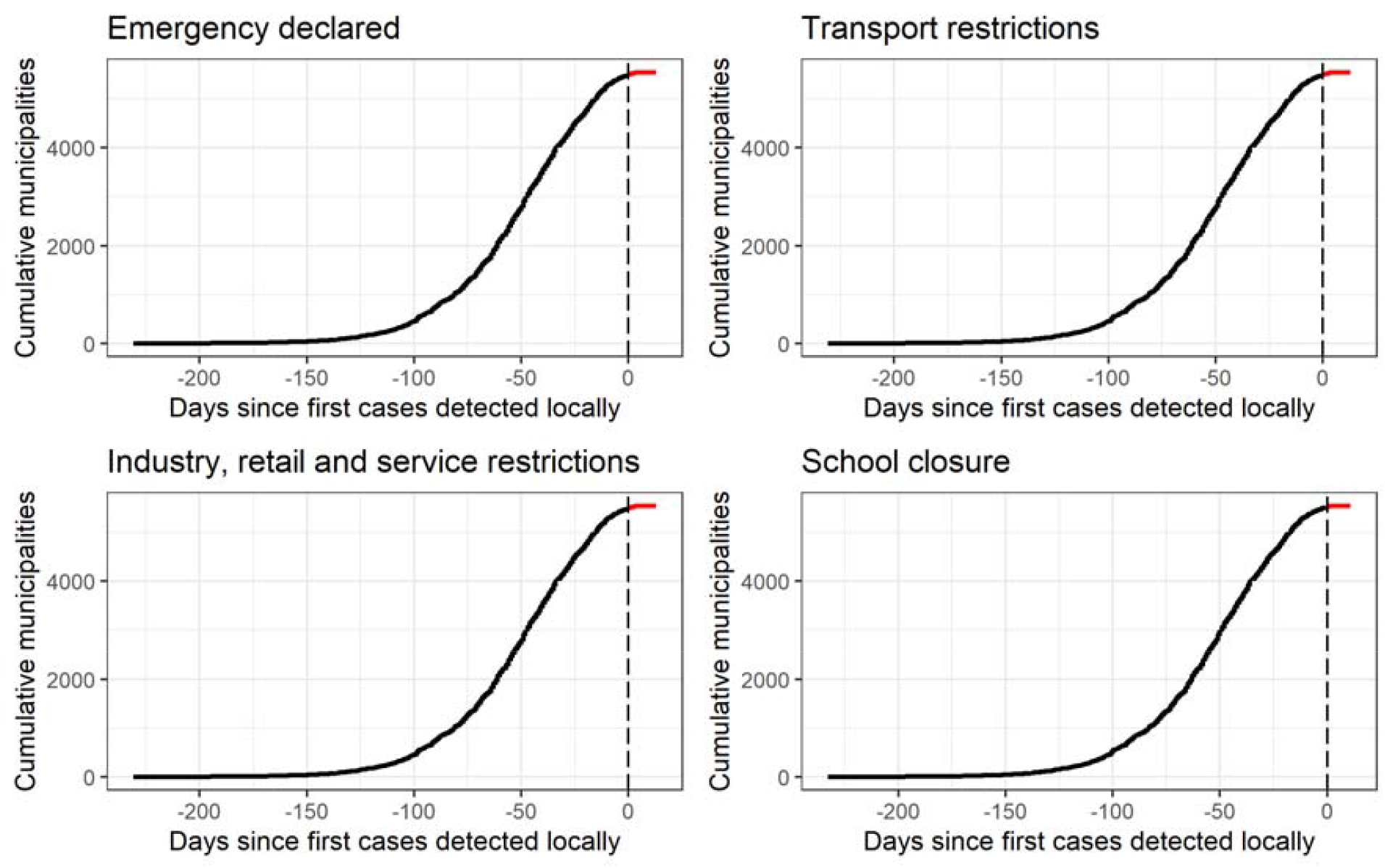
Timing of announcement of interventions relative to the first COVID-19 cases being reported locally (dashed black vertical line). Dots in black represent municipalities where restrictions were announced prior to any cases being reported, while dots in red show municipalities where restrictions were announced after cases were reported. 5,547 municipalities were included in this analysis.

### Analysis of factors associated with time for the epidemic to arrive in a municipality

Our “outbreak” threshold (standardised incidence of 10 cases per 10,000 residents) was first exceeded on April 12^th^ 2020. By the censoring date for this study of July 14^th^ 2021 all municipalities had exceeded the threshold incidence.

Consistent with the patterns of observed spread in Figures 1 and 2, the univariable analyses suggested that municipalities in the North region exceeded the outbreak threshold earlier, followed by those in the Northeast and Central-West regions and finally those in the South and Southeast regions (Table 1). After adjusting for geographic region, the epidemic can be seen to have arrived earliest in those municipalities with higher population density, higher social development index (SDI) and greater percentage of residences with piped sanitation. There was evidence that municipalities further from the main population centres in the state had a later arrival of the epidemic. Considering the magnitudes of the effect estimates, a 10% increase in the population density shortened the arrival time by 0·9 days [95% CI:0·8,0·9]. An increase of 10% in the travel time to the largest city in the State was associated with a delay in arrival of 0·4 days [95% CI:0·2,0·5]. An increase of 10% in the SDI was associated with a decrease of 11·1 days [95% CI:13·2,8·9] in the time to arrival.

**Table 1.** Tobit regression analysis of time to outbreak for each municipality (days since 31st March 2020). - Starting incidence 1 case per 10,000 residents - Epidemic onset incidence = 10 cases per 10,000 residents.

| Characteristic of the municipality |  | Frequency | Median value<br>(Interquartile range) | Unadjusted<br>(univariable) model<br>estimates<br>[95% CI] | Adjusted<br>(multivariable) model<br>estimates<br>[95% CI] |
| --- | --- | --- | --- | --- | --- |
| Geographic region | Central West | 444 |  | - | - |
|  | North | 449 |  | -32.1 [-36.7,-27.5] | -34.0 [-38.6,-29.5] |
|  | North East | 1775 |  | -15.7 [-19.4,-12.1] | -3.6 [-7.3,0.2] |
|  | South | 1157 |  | 3.7 [-0.1,7.6] | 20.8 [16.9,24.6] |
|  | South-East | 1653 |  | 4.4 [0.7,8.1] | 26.9 [23.1,30.8] |
| Population density (log<sub>n</sub>)<br>(population/km<sup>2</sup>) |  |  | 3.20<br>(2.45 - 3.96) | -5.8 [-6.5,-5.2] | -8.5 [-9.2,-7.7] |
| Percentage of residences with<br>piped water |  |  | 72.30<br>(56.29 - 84.59) | 0.0 [-0.1,0.0] | 0.0 [-0.1,0.0] |
| Percentage of residences with<br>piped sewage or septic tanks |  |  | 37.70<br>(12.75 - 70.25) | 0.0 [0.0,0.0] | -0.2 [-0.2,-0.1] |
| Travel time (log<sub>n</sub>) by road to<br>most populous municipality in<br>the state (hours) |  |  | 2.96<br>(2.34 - 3.44) | -1.0 [-2.5,0.5] | 3.6 [2.2,5.1] |
| Social Development Index<br>(SDI) |  |  | 0.25<br>(0.22 - 0.27) | -140.1 [-163.4,-<br>116.7] | -111.1 [-132.8,-89.3] |

Sensitivity analyses were carried out to investigate the effect of changing the threshold incidence for epidemic arrival from 5 to 15 cases per 10,000 residents (Tables S1 and S2 Supplementary Materials). The interpretation of the direction of the effect of the included covariates remained the same within this range, whilst there were variations in the magnitude of the effect (Table S3 Supplementary Materials).

^1^ As model contains an interaction between geographic region and start day the effect estimates for the interaction terms are shown in the expanded table below

### Analysis of factors associated with the rate of growth in the early stages of the epidemic

To measure the intensity of the epidemic in each municipality after arrival we calculated the mean reproduction number (*R*_*t*_) over the early phase of the epidemic. A total of 2757 municipalities contained sufficient data for *R*_*t*_ calculation (i.e. at least 30 days of data from the first case report and more than 200 cumulative cases) with the inter-quartile range for the estimated values ranging from 0·852 to 1·094.

From the unadjusted analyses (Table 2), it can be observed that the early epidemic was more intense on average in all other regions than the Central West, with regression coefficients indicating that mean *R*_*t*_ ranged from an increase by a factor of 1·46 in the North region to 1·30 in the Southern region. Epidemic intensity decreased in the two latter time periods compared to the first, though the effect was significantly smaller than that for geographic region; the regression coefficients for the latter two time periods indicated that the effect of time was to decrease mean *R*_*t*_ by factors of 0·86 and 0·68 respectively. In the multivariable model, a high degree of heterogeneity across space and time in mean *R*_*t*_ was seen (Fig 4). In the Central West region mean *R*_*t*_ decreased from 1·02 to 0·61, over the three time periods. In the North the comparative decrease was from 1·13 to 0·72 and in the South-East from 0·77 to 0·54. There was no statistical evidence for a change in the North-East or South regions.

**Table 2.** Summary of unadjusted and adjusted multivariate mean *Rt* linear regression model. Mean *Rt* is calculated over a window from 30 to 150 days since the arrival (standardised incidence > 1 case per 10,000 residents) of COVID-19 in the respective municipality.

| Characteristic of the municipality |  | Median value<br>(Interquartile<br>range) | Freq | Unadjusted<br>(univariable)<br>model estimates<br>(95% CI) | Adjusted<br>(multivariable)<br>model estimates<br>(95% CI) |
| --- | --- | --- | --- | --- | --- |
| Geographic<br>Region | Central West |  | 228 | 1 | -<sup>1</sup> |
|  | North |  | 314 | 1.464 [1.288,1.665] | -<sup>1</sup> |
|  | North East |  | 993 | 1.328 [1.191,1.48] | -<sup>1</sup> |
|  | South |  | 470 | 1.301 [1.155,1.465] | -<sup>1</sup> |
|  | South-East |  | 752 | 1.323 [1.184,1.48] | -<sup>1</sup> |
| Date of local<br>epidemic start<br>(standardised<br>incidence > 1<br>case per<br>10,000<br>residents) | 14-Mar to 1-May<br>2020 |  | 875 | 1 | -<sup>1</sup> |
|  | 02-May to 21-May<br>2020 |  | 913 | 0.859 [0.802,0.920] | -<sup>1</sup> |
|  | 22-May to 6-Nov<br>2020 |  | 969 | 0.675 [0.631,0.723] | -<sup>1</sup> |
| Population density (log<sub>n</sub>)<br>(population/km<sup>2</sup>) |  | 3.31 (2.52 -<br>4.21) |  | 1.096 [1.078,1.116] | 1.067 [1.042,1.092] |
| Percentage of residences with<br>piped water |  | 72.76 (56.12<br>- 85.20) |  | 1.004 [1.003,1.005] | 1.003 [1.001,1.004] |
| Percentage of residences with<br>piped sewage or septic tanks |  | 36.71 (12.45<br>- 70.80) |  | 1.003 [1.002,1.004] | 1.003 [1.002,1.005] |
| Travel time (log<sub>n</sub>) by road to most<br>populous municipality in the state<br>(hours) |  | 2.96 (2.32 -<br>3.46) |  | 1.022 [0.981,1.065] | 1.012 [0.966,1.062] |
| Social Development Index (SDI) |  | 0.25 (0.22 -<br>0.28) |  | 3.834 [1.978,7.441] | 3.300 [1.652,6.593] |

<sup>1</sup> As model contains an interaction between geographic region and start day the effect estimates for the interaction terms are shown in the expanded table below

**Figure 4.**
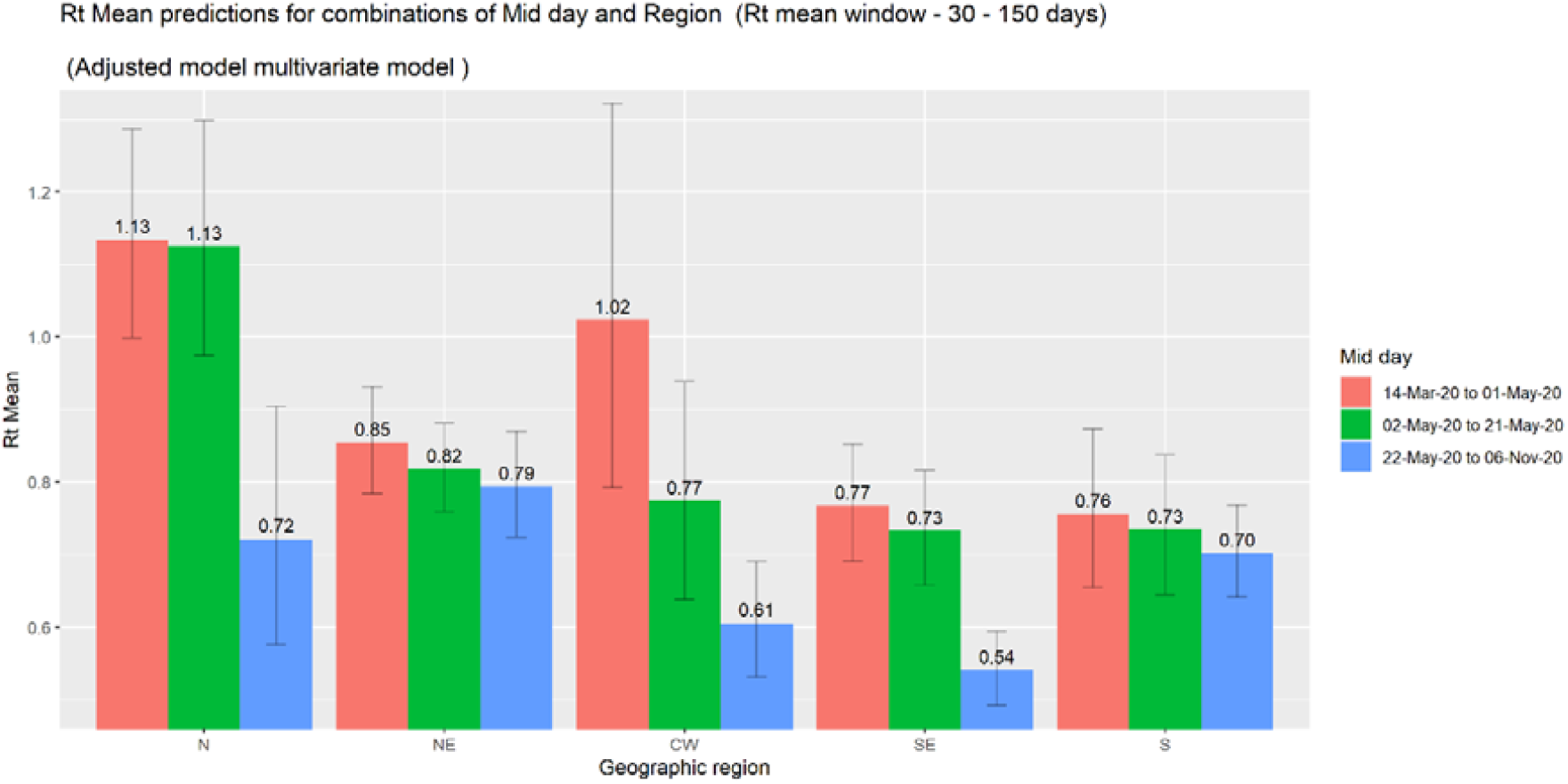
Estimated marginal mean values of *Rt* mean in the final adjusted multivariate linear regression model^*^. ^*^ The Codes for geographic region are ; North (N), Northeast (NE), Central-West (CW), Southeast (SE) and South (S) see map in Figure1.

The effects on mean *R*_*t*_ of covariates other than geographic region and time period were relatively small indicating that a large amount of the variation was not explained by these factors. From the univariable analysis it was seen that municipalities that were more densely populated, with higher levels of provision of piped water and sanitation or a higher SDI had higher mean *R*_*t*_ values (Table 2), whilst those further from the main population centre in the state had lower values. In the adjusted multivariable analysis these associations were retained with marginally lower effect estimates. There was no evidence that mean *R*_*t*_ was associated with the travel time to the most populous municipality in the state (adjusted coefficient = 1·02 [95% CI 0·966,1·062]

A sensitivity analysis was carried out in which the end date for the calculation of mean *R*_*t*_ was adjusted over a range from 100 to 180 days. The unadjusted and adjusted models for the extreme values are presented in Tables S4 and S5 in the Supplementary Materials. Whilst there were small changes for the effect estimates the trends in the associations see for the 30 to 150 day range remained unchanged, increasing the strength of evidence for the findings reported.

### Predicting new maximum values of incidence

Figure 5 shows the values of the area under the ROC curve (AUC) for the ability to predict a new maximum incidence in the following 30 days. The values are generally between 0·70 and 0·90, corresponding to accuracy described previously as “useful for some purposes”^45^. For the Central West region, AUCs are high (between 0·8 and 0·9) after a spike in incidence in late 2020, indicating that the lack of subsequent peaks was predictable. Overall, the AUC values are associated with incidence, suggesting that the method is better able to learn across municipalities when higher or lower rates are propagating across the country, i.e. that increases elsewhere helped predict peaks in each index municipality. From each ROC curve, values of sensitivity and specificity were chosen to maximize the sum of these two parameters. For sensitivity, averaging over time, the regions had similar values, between 70 and 73%. For specificity, the average values ranged from 63% for the South-East region to 74% in the North.

**Figure 5.**
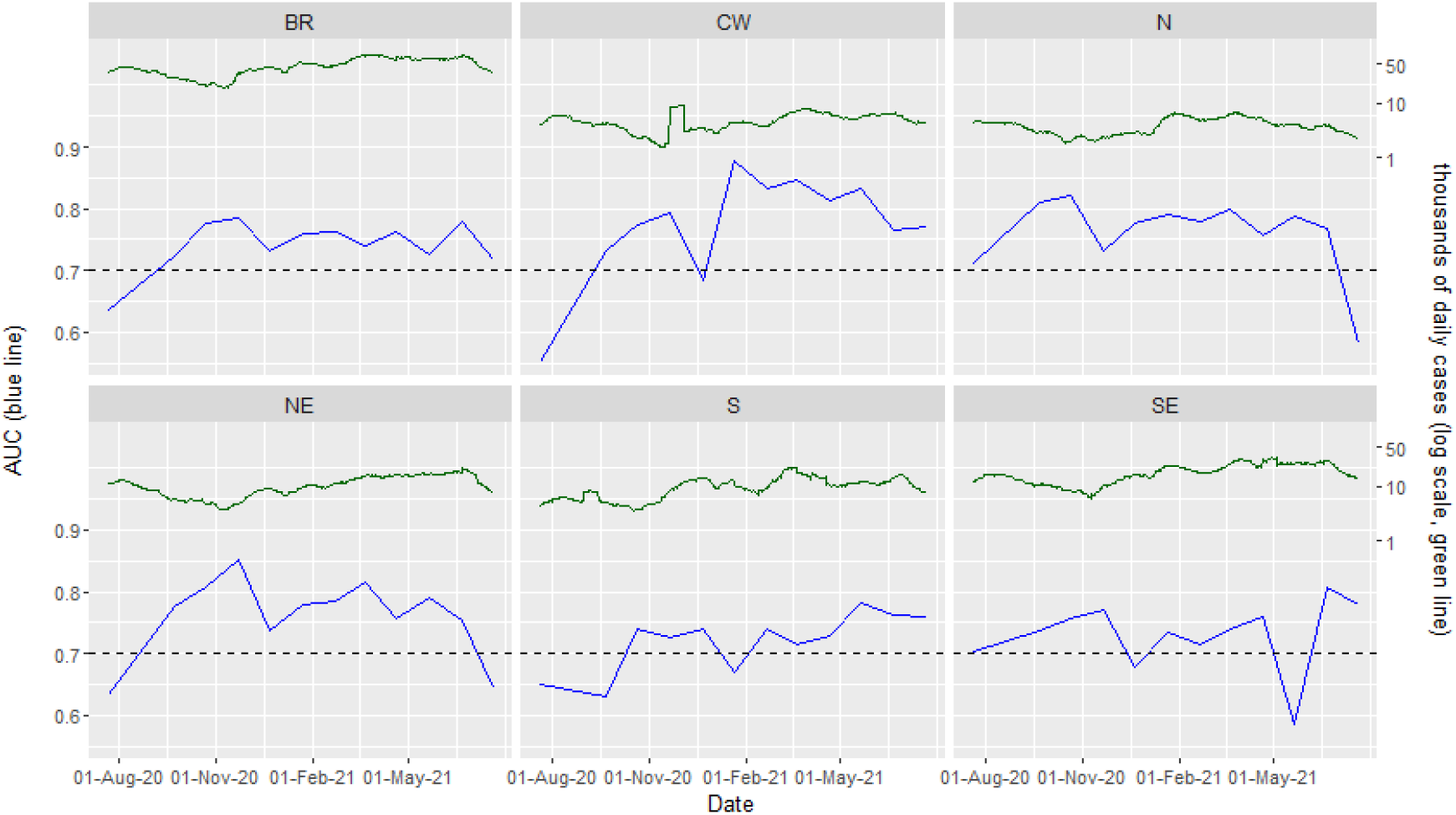
Area under the curve (AUC) for prediction of a new maximum incidence in the following 30 days (left vertical axis, blue line) against calendar time (horizontal axis). The daily number of incident cases (rolling fortnightly average) is shown in the green line, right axis. The dashed line shows the lower end of the range 0·70-0·90. There is one panel for each region (Central-West, North, North-East, South and South-East), with the “BR” panel showing all Brazil.

## Discussion

We describe the development of an online application, CLIC Brazil, that allows comparison of the spread and impact of the COVID-19 epidemic in Brazil between local areas (municipalities). We show how basic analyses, largely available through the application can be used to identify pathways and determinants of spread. Further we identify and explain heterogeneities in burden and assess the relative timeliness of reactive interventions. Underlying the app is a portable data processing and analysis pipeline which enables real-time comparisons of spatially disaggregated COVID-19 epidemic trajectories over time. The technical framework described is modular and generalisable and could be used for monitoring future disease epidemics, provided that real-time geographically located surveillance data is available.

Our analyses show that despite an initial identification of SARS Cov-2 in the large South Eastern cities of São Paulo and Rio de Janeiro^46,47^ the early focus of the epidemic quickly shifted to the Northern region before spreading to North Eastern coastal cities and then to the Southern and South Eastern regions of the country. This was then followed by a resurgence of transmission and higher levels of incidence in the North linked temporally to the emergence of the Gamma (P1) variant in the same area. Subsequently these higher rates of infection have been seen in most areas of the country and remain high currently (August 2021).

It might have been expected that individuals resident in wealthier areas would have experienced a less serious impact of the epidemic due to having better access to healthcare for those with serious illness and being more likely to be able to adopt social distancing measures designed to mitigate infection. Our findings were in contrast to this and suggest that in general, places with higher social development indices (SDI) experienced an earlier arrival and more rapid early propagation of the epidemic. This finding may be due in part to greater provision of and access to testing in areas with higher SDI, particularly given the greater role private sector testing played in the earlier stages of the epidemic. Also, the covariates used in the derivation of the social development index may not fully reflect the impact of wealth and employment type or the ability of individuals in different areas to adopt social distancing and lessen their risk of infection.

A study using data aggregated at the country level from the 5 BRICS countries (Brazil, Russia, China, India and South Africa) showed that COVID-19 case numbers were associated with increasing levels of poverty^48^. Other studies have shown that COVID-19 mortality rates were greater in areas of Brazil with lower levels of various socio-economic indicators ^49,50^. Possible explanations for the differences with our findings are that we focussed on the early stages of the local epidemic (time to arrival and early rate of propagation) and also that we were comparing trends between municipalities within one country. Also we used COVID-19 case incidence rather than mortality as our outcome, it would be expected that those in areas with higher levels of poverty experience greater barriers to accessing adequate healthcare and hence be likely to experience more severe COVID-19 disease outcomes.

Places that were more distant from major population centres experienced later onset though there was no evidence that the propagation was slower once the infection became established locally. The delayed arrival may be related to transport connectivity, however weaker health infrastructure and lower provision of testing may also be relevant.

There were large differences in the mean *Rt* between local areas and over time that could not be explained by the covariates included in our analysis, These differences may be related to differences in patterns of social mixing or the imposition of, and the level of adherence to, non-pharmaceutical interventions (NPIs), and the extent to which the roll-out of public measure designed to curb the spread of the epidemic was devolved to a local level by national and State authorities ^51^.

There was evidence, at least for the Central-West, North and South-East regions, that the reproduction number was lower in places where the epidemic arrived later. Studies have shown that in general individuals are more likely to change their health behaviour if they perceive they are at a heightened level of risk ^52^ Hence it may be hypothesised that individuals were more likely to adhere to social distancing and other NPIs as their awareness of the severity of COVID-19 infection grew. A recent worldwide assessment of changes in adherence to NPI’s to mitigate COVID-19 indicated that whilst this may be true for interventions with a low economic cost, such as mask wearing, it was not the case for adherence to social isolation which had a higher economic cost ^53^. More work on adherence to NPIs in Brazil is urgently needed given the unique socio-political approach the country took to COVID-19 control, particularly in the early stages of the epidemic.

The algorithm developed to predict the likelihood of a place experiencing a new record level of incidence in the following 30 days showed reasonably good predictive values once the epidemic had become established in each region and nationally. This suggests that the approach would be useful for use assessing the immediate local impact of measures taken to mitigate the COVID-19 epidemic.

### Limitations

There are several limitations of this study that should be acknowledged. It is likely that the consistency of reporting of COVID-19 cases by state health departments differed both between places and over time. Also, the travel time covariate, a measure of geographical isolation, estimates the time for within state journeys to the most populous city in the state. Those living on the border region of a state may be distant from the most populous city in that state but closer to a large city in a neighbouring state, however, given the small effect sizes for the association with travel time the effect on the outcome would be small. The stratification of municipalities into five broad regions whilst providing a reasonable number of strata for the analyses was not able to account for within region variation in geographically associated characteristics. Additionally we recognise that as a clearer understanding of the determinants of COVID-19 infection in the Brazilian population becomes available, future studies comparing disease incidence between different areas should include standardisation by a wider range of risk factors possibly including gender, co-morbidities and race

Since December 2020 cases have begun to increase again, initially focussed on cities such as Manaus in the Northern Amazonas ^3,54,55^.There is evidence this is driven by the local emergence of a new variant of concern, Gamma (P1). which has higher transmissibility and exhibits the ability to evade the neutralising effect of antibodies to previous infection ^54^. The Gamma (P1) variant has rapidly spread to become the dominant strain in Amazonas state and throughout the country ^55^. Analyses of factors associated with early development of the epidemic may be useful in predicting the spread of this new variant. Continual tracking of this next phase of the epidemic using methods demonstrated in our analyses will be important to assess similarities and differences in the spatial spread of different COVID-19 epidemic waves. These analyses should include spatially disaggregated data on vaccine coverage ^9^ to assess the impact of the epidemic on vaccination on the development of the epidemic. Pairing these outputs with phylogenetic analyses ^3,54^ of SARS-CoV-2 virus samples would enable a more detailed picture of past and present subnational spread of the epidemic.

## Conclusion

This study demonstrates that by monitoring, standardising and analysing the development of an epidemic at a local level, insights can be gained into spatial and temporal heterogeneities. Such insights are often impossible using raw case counts or when data are aggregated over larger areas. We show the utility of using age-standardised incidence as a comparable epidemiological metric for a variety of analyses and have developed an on-line application that allows a range of stakeholders to simply compare and contrast the evolution of the COVID-19 epidemic in different areas. This approach could prove useful for real-time local monitoring and analysis of a range of other emerging infectious disease outbreaks.

## Supporting information

Screen shots

Sensitivity analyses

## Data Availability

All code described in the paper can be downloaded from this github repository - https://github.com/Paul-Mee/clic_brazil . Data may be downloaded from the public domain source Brazil.io

## Acknowledgements

This project was supported by a Medical Research Council UK (MRC-UK) and the São Paulo Research Foundation (FAPESP) Newton partnership award (MR/S0195/1 and FAPESP 18/14389-0) for the CADDE project (http://caddecentre.org/). N.R.F. is supported by a Wellcome Trust and Royal Society Sir Henry Dale Fellowship: (204311/Z/16/Z). OJB was supported by a Wellcome Trust Sir Henry Wellcome Fellowship (206471/Z/17/Z). CAPJ was supported by FAPESP (2019/21858-0), Fundação Faculdade de Medicina and Coordenação de Aperfeiçoamento de Pessoal de Nível Superior – Brasil (CAPES) – Finance Code 001.

*The following authors were part of the Centre for Mathematical Modelling of Infectious Disease COVID-19 working group. Each contributed in processing, cleaning and interpretation of data, interpreted findings, contributed to the manuscript, and approved the work for publication: Naomi R Waterlow, C Julian Villabona-Arenas, James D Munday, Graham F. Medley, Rachel Lowe, Yang Liu, Amy Gimma, Kevin van Zandvoort, Joel Hellewell, Damien C Tully, Megan Auzenbergs, Gwenan M Knight, Adam J Kucharski, Rosanna C Barnard, William Waites, W John Edmunds, Nikos I Bosse, Akira Endo, Emilie Finch, Timothy W Russell, Yung-Wai Desmond Chan, Matthew Quaife, Rosalind M Eggo, Kiesha Prem, Rachael Pung, Thibaut Jombart, Billy J Quilty, Samuel Clifford, Mihaly Koltai, Hamish P Gibbs, Sam Abbott, Christopher I Jarvis, Yalda Jafari, Petra Klepac, Fabienne Krauer, Fiona Yueqian Sun, Sebastian Funk, Frank G Sandmann, Emily S Nightingale, Jiayao Lei, Sophie R Meakin, Alicia Rosello, Carl A B Pearson, David Hodgson, Ciara V McCarthy, Anna M Foss, Katherine E. Atkins*,

## References

1. WHO Coronavirus (COVID-19) Dashboard | WHO Coronavirus Disease (COVID-19) Dashboard. Accessed March 24, 2021. https://covid19.who.int/

2. Castro MC, Kim S, Barberia L, et al. Spatiotemporal pattern of COVID-19 spread in Brazil. Science (80-). Published online April 14, 2021:eabh1558. doi:10.1126/science.abh1558

3. Faria NR, Mellan TA, Whittaker C, et al. Genomics and epidemiology of the P.1 SARS-CoV-2 lineage in Manaus, Brazil. Science (80-). Published online April 14, 2021:eabh2644. doi:10.1126/science.abh2644

4. Dowd JB, Andriano L, Brazel DM, et al. Demographic science aids in understanding the spread and fatality rates of COVID-19. Proc Natl Acad Sci U S A. 2020;117(18):9696–9698. doi:10.1073/pnas.2004911117

5. Dudel C, Riffe T, Acosta E, van Raalte A, Strozza C, Myrskylä M. Monitoring trends and differences in COVID-19 case-fatality rates using decomposition methods: Contributions of age structure and age-specific fatality. Masquelier B, ed. PLoS One. 2020;15(9):e0238904. doi:10.1371/journal.pone.0238904

6. Ferrante L, Steinmetz WA, Almeida ACL, et al. Brazil’s policies condemn Amazonia to a second wave of COVID-19. Nat Med. 2020;26(9):1315. doi:10.1038/s41591-020-1026-x

7. Hallal PC, Hartwig FP, Horta BL, et al. SARS-CoV-2 antibody prevalence in Brazil: results from two successive nationwide serological household surveys. Lancet Glob Heal. 2020;8(11):e1390–e1398. doi:10.1016/S2214-109X(20)30387-9

8. de Souza WM, Buss LF, Candido D da S, et al. Epidemiological and clinical characteristics of the COVID-19 epidemic in Brazil. Nat Hum Behav. 2020;4(8):856–865. doi:10.1038/s41562-020-0928-4

9. Painel da vacinação COVID-19. Accessed August 20, 2021. https://apps.kauebraga.dev/shiny/painel_vacinacao_covid/

10. Jombart T, Ghozzi S, Schumacher D, et al. Real-time monitoring of COVID-19 dynamics using automated trend fitting and anomaly detection. medRxiv. Published online September 3, 2020:2020.09.02.20186502. doi:10.1101/2020.09.02.20186502

11. COVID-19 Map - Johns Hopkins Coronavirus Resource Center. Accessed August 20, 2021. https://coronavirus.jhu.edu/map.html

12. Dong E, Du H, Gardner L. An interactive web-based dashboard to track COVID-19 in real time. Lancet Infect Dis. 2020;20(5):533–534. doi:10.1016/S1473-3099(20)30120-1

13. COVID-19 ITALIA - Desktop. Accessed August 20, 2021. https://opendatadpc.maps.arcgis.com/apps/opsdashboard/index.html#/b0c68bce2cce478ea ac82fe38d4138b1

14. EpiForecasts - We forecast infectious disease outbreaks. Accessed August 20, 2021. https://epiforecasts.io/

15. Abbott S, Hellewell J, Thompson RN, et al. Estimating the time-varying reproduction number of SARS-CoV-2 using national and subnational case counts. Wellcome Open Res. 2020;5:112. doi:10.12688/wellcomeopenres.16006.1

16. CDC COVID Data Tracker. Accessed August 20, 2021. https://covid.cdc.gov/covid-data-tracker/#forecasting_weeklydeaths

17. Ray EL, Wattanachit N, Niemi J, et al. Ensemble forecasts of Coronavirus Disease 2019 (COVID-19) in the U.S. medRxiv. Published online August 22, 2020:2020.08.19.20177493. doi:10.1101/2020.08.19.20177493

18. Cramer EY, Ray EL, Lopez VK, et al. Evaluation of individual and ensemble probabilistic forecasts of COVID-19 mortality in the US. medRxiv. 2021;46:2021.02.03.21250974. doi:10.1101/2021.02.03.21250974

19. Short-term forecasts of COVID-19 deaths in multiple countries. Accessed April 29, 2021. https://mrc-ide.github.io/covid19-short-term-forecasts/

20. Gostic KM, McGough L, Baskerville EB, et al. Practical considerations for measuring the effective reproductive number, Rt. Pitzer VE, ed. PLOS Comput Biol. 2020;16(12):e1008409. doi:10.1371/journal.pcbi.1008409

21. Cori A, Ferguson NM, Fraser C, Cauchemez S. A New Framework and Software to Estimate Time-Varying Reproduction Numbers During Epidemics. Am J Epidemiol. 2013;178(9):1505–1512. doi:10.1093/aje/kwt133

22. Cauchemez S, Boëlle PY, Thomas G, Valleron AJ. Estimating in real time the efficacy of measures to control emerging communicable diseases. Am J Epidemiol. 2006;164(6):591–597. doi:10.1093/aje/kwj274

23. Covid-19: National and Subnational estimates for Brazil. Accessed March 9, 2021. https://epiforecasts.io/covid/posts/national/brazil/

24. World Population Prospects - Population Division - United Nations. Accessed August 3, 2021. https://population.un.org/wpp/

25. Brazil.IO. Brazil.IO: COVID-19 Source: Health Departments of the Federative Units, data process. https://brasil.io/dataset/covid19/

26. Instituto Brasileiro de Geografia e Estatisitica (IBGE). IBGE - Census 2010. https://www.ibge.gov.br/en/statistics/social/income-expenditure-and-consumption/18391-2010-population-census.html?edicao=19720&t=publicacoes

27. WorldPop. Accessed August 20, 2021. https://www.worldpop.org/

28. Sorichetta A, Hornby GM, Stevens FR, Gaughan AE, Linard C, Tatem AJ. High-resolution gridded population datasets for Latin America and the Caribbean in 2010, 2015, and 2020. Sci Data. 2015;2(1):1–12. doi:10.1038/sdata.2015.45

29. Kraemer MUG, Sadilek A, Zhang Q, et al. Mapping global variation in human mobility. Nat Hum Behav. 2020;4(8):800–810. doi:10.1038/s41562-020-0875-0

30. Pfeffer DA, Lucas TCD, May D, et al. MalariaAtlas: An R interface to global malariometric data hosted by the Malaria Atlas Project. Malar J. 2018;17(1):352. doi:10.1186/s12936-018-2500-5

31. Wang H, Naghavi M, Allen C, et al. Global, regional, and national life expectancy, all-cause mortality, and cause-specific mortality for 249 causes of death, 1980–2015: a systematic analysis for the Global Burden of Disease Study 2015. Lancet. 2016;388(10053):1459–1544. doi:10.1016/S0140-6736(16)31012-1

32. Pereira, R.H.M; Gonçalves CN. et al. geobr: Loads Shapefiles of Official Spatial Data Sets of Brazil. GitHub repository. Published 2019. https://github.com/ipeaGIT/geobr.

33. Chang W, Cheng J, Allaire JJ, et al. shiny: Web Application Framework for R. Published online 2021. https://cran.r-project.org/package=shiny

34. Wickham H. Ggplot2: Elegant Graphics for Data Analysis. Springer-Verlag New York; 2016. https://ggplot2.tidyverse.org

35. Cheng J, Karambelkar B, Xie Y. leaflet: Create Interactive Web Maps with the JavaScript “Leaflet” Library. Published online 2021. https://cran.r-project.org/package=leaflet

36. Parag KV. Improved estimation of time-varying reproduction numbers at low case incidence and between epidemic waves. medRxiv. Published online September 18, 2020:2020.09.14.20194589. doi:10.1101/2020.09.14.20194589

37. Ferguson NM. Report 9 - Impact of non-pharmaceutical interventions (NPIs) to reduce COVID-19 mortality and healthcare demand | Faculty of Medicine | Imperial College London. Accessed April 22, 2021. https://www.imperial.ac.uk/mrc-global-infectious-disease-analysis/covid-19/report-9-impact-of-npis-on-covid-19/

38. Lima FET, Albuquerque NLS de, Florencio S de SG, et al. Intervalo de tempo decorrido entre o início dos sintomas e a realização do exame para COVID-19 nas capitais brasileiras, agosto de 2020*. Epidemiol e Serviços Saúde. 2021;30(1):e2020788. doi:10.1590/s1679-4974202100010002

39. Parag K V., Donnelly CA. Using information theory to optimise epidemic models for real-time prediction and estimation. Ferrari M (Matt), ed. PLOS Comput Biol. 2020;16(7):e1007990. doi:10.1371/journal.pcbi.1007990

40. Tobin J. Estimation of Relationships for Limited Dependent Variables. Econometrica. 1958;26(1):24. doi:10.2307/1907382

41. VGAM. Accessed April 29, 2021. https://www.stat.auckland.ac.nz/~yee/VGAM/

42. GitHub - therneau/survival: Survival package for R. Accessed April 29, 2021. https://github.com/therneau/survival

43. Benchimol EI, Smeeth L, Guttmann A, et al. The REporting of studies Conducted using Observational Routinely-collected health Data (RECORD) Statement. PLOS Med. 2015;12(10):e1001885. doi:10.1371/journal.pmed.1001885

44. Candido DS, Claro IM, de Jesus JG, et al. Evolution and epidemic spread of SARS-CoV-2 in Brazil. Science (80-). 2020;369(6508):1255–1260. doi:10.1126/SCIENCE.ABD2161

45. Swets JA. Measuring the accuracy of diagnostic systems. Sci Sci. 1988;240(4857):1285–1293. doi:10.1126/science.3287615

46. de Jesus JG, Sacchi C, Candido D da S, et al. Importation and early local transmission of covid-19 in brazil, 2020. Rev Inst Med Trop Sao Paulo. 2020;62. doi:10.1590/S1678-9946202062030

47. Candido DDS, Watts A, Abade L, et al. Routes for COVID-19 importation in Brazil. J Travel Med. 2020;27(3):1–3. doi:10.1093/jtm/taaa042

48. Zhu J, Yan W, zhu L, Liu J. COVID-19 pandemic in BRICS countries and its association with socio-economic and demographic characteristics, health vulnerability, resources, and policy response. Infect Dis Poverty 2021 101. 2021;10(1):1–8. doi:10.1186/S40249-021-00881-W

49. Ribeiro H, Lima VM, Waldman EA. In the COVID-19 pandemic in Brazil, do brown lives matter? Lancet Glob Heal. 2020;8(8):e976–e977. doi:10.1016/S2214-109X(20)30314-4

50. Li SL, Pereira RHM, Jr CAP, et al. Higher risk of death from COVID-19 in low-income and non-White populations of São Paulo, Brazil. BMJ Glob Heal. 2021;6(4):e004959. doi:10.1136/BMJGH-2021-004959

51. de Souza Santos AA, Candido D da S, de Souza WM, et al. Dataset on SARS-CoV-2 non-pharmaceutical interventions in Brazilian municipalities. Sci Data. 2021;8(1):73. doi:10.1038/s41597-021-00859-1

52. Brewer NT, Chapman GB, Gibbons FX, Gerrard M, McCaul KD, Weinstein ND. Meta-analysis of the relationship between risk perception and health behavior: The example of vaccination. Heal Psychol. 2007;26(2):136–145. doi:10.1037/0278-6133.26.2.136

53. Petherick A, Goldszmidt R, Andrade EB, et al. A worldwide assessment of changes in adherence to COVID-19 protective behaviours and hypothesized pandemic fatigue. Nat Hum Behav 2021. Published online August 3, 2021:1–16. doi:10.1038/s41562-021-01181-x

54. Sabino EC, Buss LF, Carvalho MPS, et al. Resurgence of COVID-19 in Manaus, Brazil, despite high seroprevalence. Lancet. 2021;397(10273):452–455. doi:10.1016/S0140-6736(21)00183-5

55. Naveca F, Souza V, Corado A, et al. COVID-19 epidemic in the Brazilian state of Amazonas was driven by long-term persistence of endemic SARS-CoV-2 lineages and the recent emergence of the new Variant of Concern P.1. Published online February 25, 2021. doi:10.21203/rs.3.rs-275494/v1

