## Supplementary material for "Tracking the emergence of disparities in the subnational spread of COVID-19 in Brazil using an online application for real-time data visualisation: a longitudinal analysis": Screen shots

**Supplementary Materials**

**Screenshots from the CLIC Brazil app**

**Supplementary figure S3 Comparison of trends**

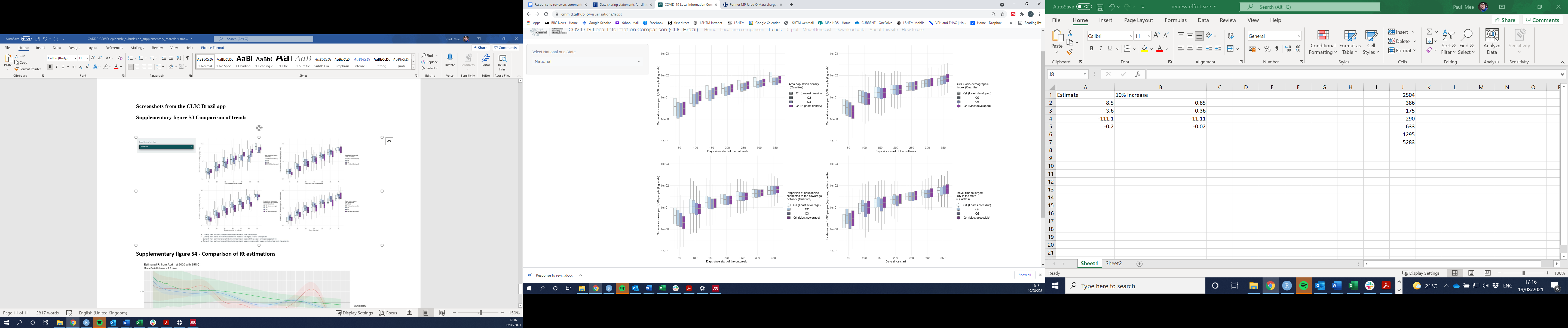

**Supplementary figure S4 - Comparison of Rt estimations**

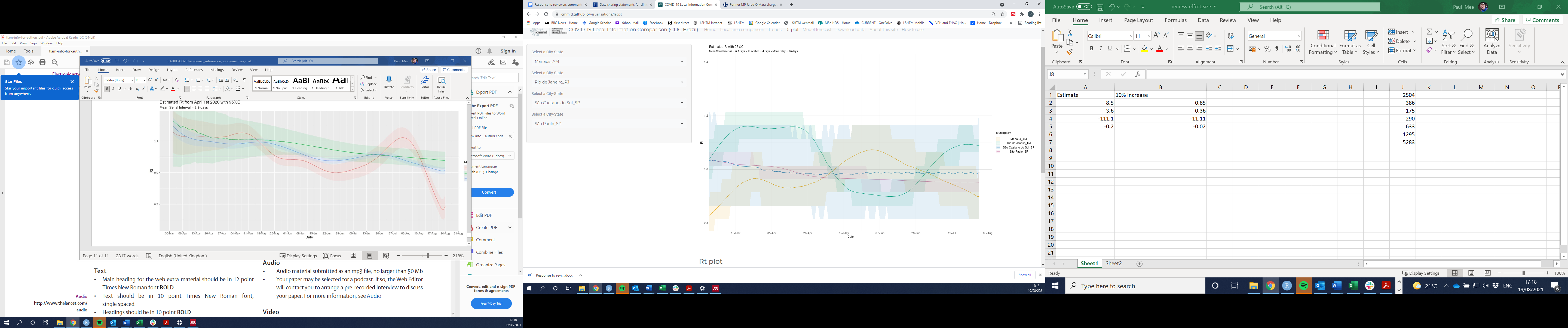

**Supplementary figure S5 - Comparison of model forecasts**

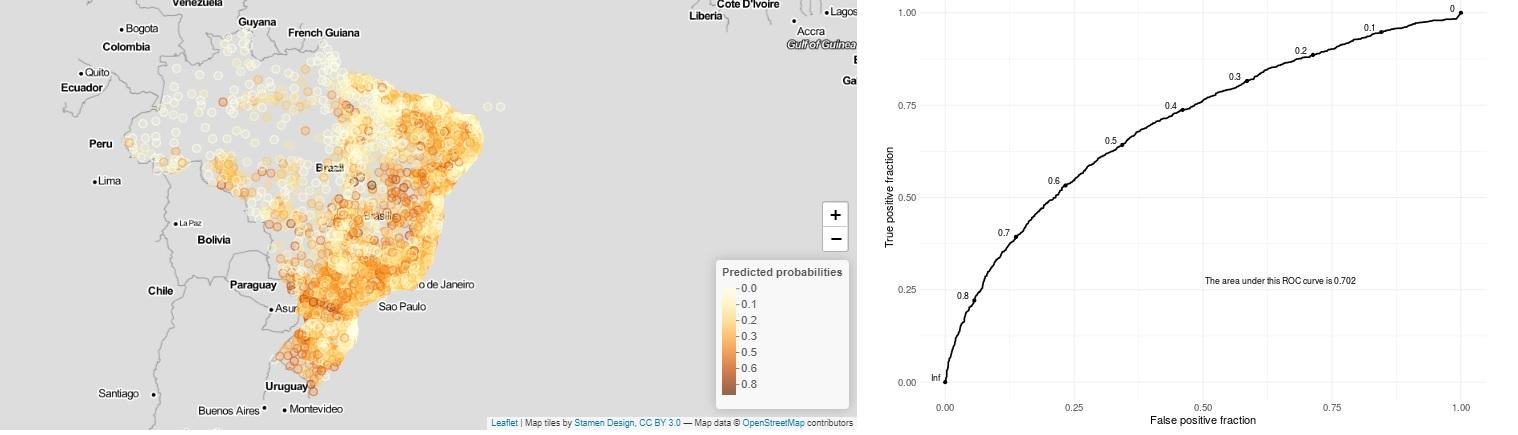
