## Supplementary methods for "Tracking the emergence of disparities in the subnational spread of COVID-19 in Brazil using an online application for real-time data visualisation: a longitudinal analysis"

**Supplementary Materials**

**Data Sources**

**T**he numbers of COVID-19 cases and deaths, aggregated by municipality were automatically downloaded daily from the Brazil.io COVID-19 project repository (1) .This repository contains data extracted from the bulletins of State Health secretariats. Data on the distribution of the population by age and the sociodemographic characteristics of each municipality were obtained from most recent national demographic census, run by the Instituto Brasileiro de Geografia e Estatística (IBGE) in 2010 (2). The extracted covariates included the mean population density (population/km^2^), the percentage of households with access to piped water, the percentage of households with access to a sewage system or septic tank, the mean per capita income, educational attainment and fertility rates.

Data on the age distribution of COVID-19 cases were derived from case reports throughout Brazil between 2^nd^ February and 25^th^ March 2020, collected by the Brazilian Ministry of Health and were used with their permission. This data from the early stage of the epidemic was subsequently compared with Severe Acute Respiratory Syndrome surveillance data collected in the SIVEP-Gripe respiratory disease surveillance system (3) . There was no evidence that the age distribution of COVID-19 cases had changed significantly over time (Supplementary data figure S2). The data were initially processed to account for typographical differences in place names between data sets. For some municipalities, the cumulative number of cases or deaths were occasionally observed to decrease, due to the inclusion of a negative daily case count. Visual inspection of the data indicated that these negative counts were often followed by a large positive count, perhaps as a data correction. To correct for negative increments, a process of ‘negative spreading’ was used, which acted as a post-hoc data filter to ensure we displayed cumulative data. Here the decrement in cases was set to zero on the day it was reported and then subtracted from the number of cases reported on the next day. This was repeated iteratively until the corrected number of incident cases for a particular day was at least zero. Out of a total of 2,105,460 daily case reports analysed there were 18,536 case decrements (0.88%) the median and minimum values were -1 cases, the maximum -1012 cases.

The travel time in hours from each municipality to the most populous metropolitan area in the state was calculated using [WorldPop](https://www.worldpop.org/) population data (4,5) and the [Malaria Atlas Project](https://developers.google.com/earth-engine/datasets/catalog/Oxford_MAP_friction_surface_2015_v1_0) travel time friction surface using the [Malaria Atlas (5)](https://malariaatlas.org/application-project/malariaatlas_package/) accumulated cost route finding algorithm within the [malariaAtlas](https://malariaatlas.org/application-project/malariaatlas_package/) R package (6,7) . The socio-demographic Index (SDI) is a composite average of the rankings of the incomes per capita, average educational attainment, and fertility rates scaled between 0 (lowest) and 1 (highest) (8) . The geographic region in which each place was located was assigned using a standardised designation which groups the States and the Federal District of Brasilia into five macro regions of national planning (9) (Figure 1)

Data on COVID-19 cases and deaths were censored on January 14^th^ 2021. Data on the types of non-pharmaceutical interventions implemented, and the dates of their announcement were extracted from data collated by the [Cepal Observatory](https://www.cepal.org/en/topics/covid-19) with edits and updates on timing of interventions at the municipality and state level by Andreza Aruska de Souza Santos (10,11)

**Data Standardisation**

First, the raw number of cases reported in each municipality are assigned an estimated age distribution based on the nationally aggregated Brazilian age distribution of COVID-19 cases (Appendix - Supplementary figure S1) . We then use local age-stratified population data from the 2010 national census (2) to calculate age-specific incidence of infection at the municipality level. All municipality-level age-specific incidence values are then applied to the same national age structure to generate a comparable measure of standardised incidence of COVID-19 cases per 1,000 inhabitants. Numerically, if *m_i,_* is the total number of COVID-19 cases in municipality *i*, and *n_a_* is the number of COVID-19 cases in age group *a* nationally then the estimated age distribution of cases in each municipality is given by

$$m_{i,a}=\frac{m_{i,+}n_{a}}{\Sigma_{a}n_{a}}.$$

Then, if *p_i_*_,_*_a_* is the population in each age group in each municipality and *c_a_* the national population in each age group then the standardised incidence (*s_i_*) per 1000 people in municipality *i* is calculated as follows:

$$s_{i}=1000\frac{\sum_{a} \left( \frac{c_{a}m_{i,a}}{p_{i,a}} \right)}{\Sigma_{a}c_{a}}.$$

**Forward and Backward stepwise variable selection**

In the forward stepwise approach variables were added and retained if the likelihood ratio test (LRT) statistic indicated that they significantly improved the model fit. In the backward stepwise method all variables were included and dropped iteratively (by LRT) to assess whether their omission worsened the model fit. The final model was selected as the ‘best’ model (by LRT) from the two selection approaches.

***R_t_* Estimation**

The raw case count data was adjusted to account for the differential probabilities of COVID-19 cases being reported on each day of the week (heaping). This was done by developing a generalised additive model (GAM) (12) as implemented in the R package mgcv (13) , using the negative binomial distribution family, with a spline function of number of days since Jan 1st 2020, and a fixed effect for day of the week. More specifically, thin plate regression splines were used, which fit the response variable (case counts) as a smooth function of the covariate (days since Jan 1st 2020). The spline function is specified in terms of a given number of basis functions, and this number was set to nine less than the number of weeks of available data as this provided adequate complexity to fit the variation in the data without excessive overfitting. To obtain a smoothed time series, predictions were made from the resultant GAM model using predictions from seven datasets based on the original one: in the first such dataset, all days of the week were set to Monday, in the second all days of the week were set to Tuesday and so on. An average of the seven predicted case counts was calculated and then smoothed over a +/-3 day window, using a moving average.

**Predicting whether a new maximum incidence will occur**

We used Cox regression as implemented in the “coxph” function in the “survival” package in R (14) to estimate the probability of each municipality surpassing its previous maximum weekly standardized incidence (i.e. a new “record” incidence) within the following four weeks . The analysis time was the number of weeks since the start of the epidemic (cumulative standardised incidence exceeded 1 case per 10,000). The event of interest was the setting of a new record incidence, which in general occurs more than once. First, weeks of local maximum incidence were identified, i.e. weeks preceded and followed by lower values. Weeks with a new global maximum (or “record”) incidence for that municipality are then a subset of the local maxima, defined by being strictly greater than all previous maxima. Unsmoothed incidence values were used, on the rationale that these would be closer to the values used locally to decide any decisions in terms of interventions.

The explanatory terms for each municipality included in the model, selected by likelihood ratio test, were: the differences between current record incidence and weekly incidences one, two and three weeks previously. Additionally, we included the state in which the municipality was located and its population density. Clustering of weekly data points within the municipality was included as a frailty term (15).To estimate the probability of a new record being set over the next 4 weeks, the expected number of times that a record will be set over the next 4 weeks was predicted using the “predict” method for “coxph”. The estimated probability of any new record being set in that time is calculated as 1-e^(-1 ×^ *^nexp^*^)^ , where *nexp* is the expected number of new records. This approach takes into account the baseline hazard.

The utility of this estimation was assessed in a training dataset obtained by removing the last 4 weeks’ data. A Receiver Operating Characteristic (ROC) curve was then obtained from the predicted probabilities and the observed values (whether or not a new “record” was set) and the Area under the ROC Curve (AUC) value used as a metric for the accuracy of the estimation. In addition, from each AUC, the optimal values of sensitivity and specificity were chosen on the criterion of maximizing their sum, i.e. values which give the Youden index (16).

References

1.	Brazil.IO. Brazil.IO: COVID-19 Source: Health Departments of the Federative Units, data process [Internet]. Available from: https://brasil.io/dataset/covid19/

2.	Instituto Brasileiro de Geografia e Estatisitica (IBGE). IBGE - Census 2010 [Internet]. Available from: https://sidra.ibge.gov.br/tabela/3107

3.	Bastos LS, Niquini RP, Lana RM, Villela DAM, Cruz OG, Coelho FC, et al. COVID-19 e hospitalizações por SRAG no Brasil: uma comparação até a 12^a^ semana epidemiológica de 2020. Cad Saude Publica [Internet]. 2020 [cited 2020 Apr 28];36(4). Available from: http://www.scielo.br/scielo.php?script=sci_arttext&pid=S0102-311X2020000406001&tlng=pt

4.	WorldPop [Internet]. [cited 2021 Apr 29]. Available from: https://www.worldpop.org/

5.	Sorichetta A, Hornby GM, Stevens FR, Gaughan AE, Linard C, Tatem AJ. High-resolution gridded population datasets for Latin America and the Caribbean in 2010, 2015, and 2020. Sci Data [Internet]. 2015 Sep 1 [cited 2021 Apr 21];2(1):1–12. Available from: www.worldpop.org

6.	Kraemer MUG, Sadilek A, Zhang Q, Marchal NA, Tuli G, Cohn EL, et al. Mapping global variation in human mobility. Nat Hum Behav [Internet]. 2020 Aug 1 [cited 2021 Jan 25];4(8):800–10. Available from: https://www.nature.com/articles/s41562-020-0875-0

7.	Pfeffer DA, Lucas TCD, May D, Harris J, Rozier J, Twohig KA, et al. MalariaAtlas: An R interface to global malariometric data hosted by the Malaria Atlas Project. Malar J [Internet]. 2018 Oct 5 [cited 2021 Apr 21];17(1):352. Available from: https://malariajournal.biomedcentral.com/articles/10.1186/s12936-018-2500-5

8.	Wang H, Naghavi M, Allen C, Barber RM, Carter A, Casey DC, et al. Global, regional, and national life expectancy, all-cause mortality, and cause-specific mortality for 249 causes of death, 1980–2015: a systematic analysis for the Global Burden of Disease Study 2015. Lancet. 2016 Oct 8;388(10053):1459–544.

9.	Pereira, R.H.M; Gonçalves CN. et. al. geobr: Loads Shapefiles of Official Spatial Data Sets of Brazil. GitHub repository [Internet]. 2019. Available from: https://github.com/ipeaGIT/geobr.

10.	Faria NR, Mellan TA, Whittaker C, Claro IM, Candido D da S, Mishra S, et al. Genomics and epidemiology of the P.1 SARS-CoV-2 lineage in Manaus, Brazil. Science (80- ) [Internet]. 2021 Apr 14 [cited 2021 Apr 28];eabh2644. Available from: https://www.sciencemag.org/lookup/doi/10.1126/science.abh2644

11.	de Souza WM, Buss LF, Candido D da S, Carrera JP, Li S, Zarebski AE, et al. Epidemiological and clinical characteristics of the COVID-19 epidemic in Brazil. Nat Hum Behav [Internet]. 2020 Aug 1 [cited 2020 Sep 7];4(8):856–65. Available from: https://doi.org/10.1038/s41562-020-0928-4

12.	Wood S. Generalized Additive Models: An Introduction with R. 2nd ed. CRC Press. 2017.

13.	CRAN - Package mgcv [Internet]. [cited 2021 Apr 30]. Available from: https://cran.r-project.org/web/packages/mgcv/index.html

14.	GitHub - therneau/survival: Survival package for R [Internet]. [cited 2021 Apr 29]. Available from: https://github.com/therneau/survival

15.	Govindarajulu US, Lin H, Lunetta KL, D’Agostino RB. Frailty models: Applications to biomedical and genetic studies. Stat Med [Internet]. 2011 Sep 30 [cited 2021 Mar 10];30(22):2754–64. Available from: http://doi.wiley.com/10.1002/sim.4277

16.	Akobeng AK. Understanding diagnostic tests 3: receiver operating characteristic curves. Acta Paediatr [Internet]. 2007 May 1 [cited 2021 Apr 28];96(5):644–7. Available from: http://doi.wiley.com/10.1111/j.1651-2227.2006.00178.x

**Supplementary figure S1 - Estimated percentage of all COVID-19 cases in each 5 year age group for those aged less than 90 years based on the age population distribution and COVID case distributions for Brazil.**

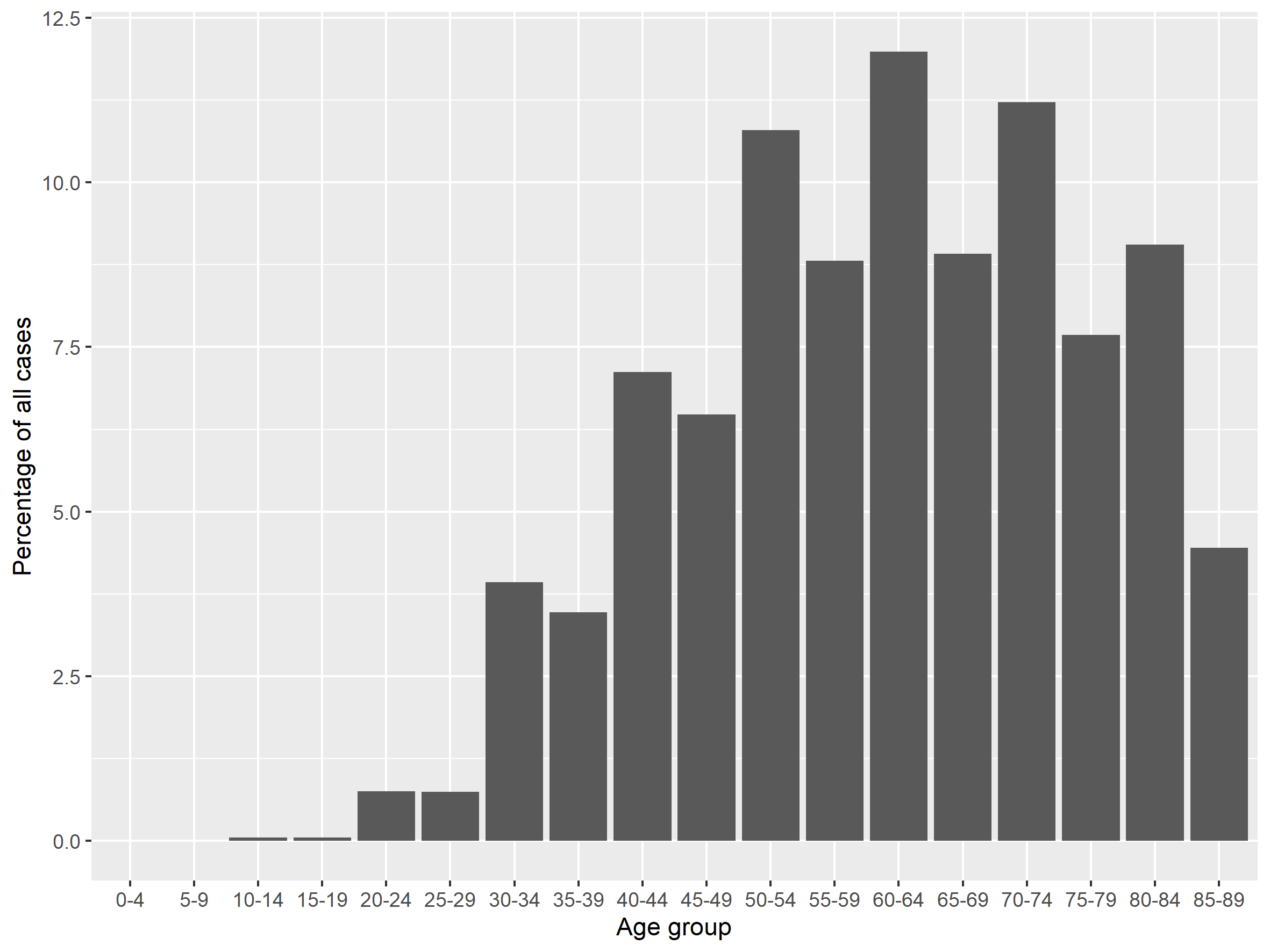

**Supplementary figure S2 – Comparison of the age distributions of COVID-19 cases from the Brazilian Ministry of Health (MoH) data for Feb to March 2020 and Severe Acute Respiratory System surveillance (SARS) data in the SIVEP-Gripe data system**

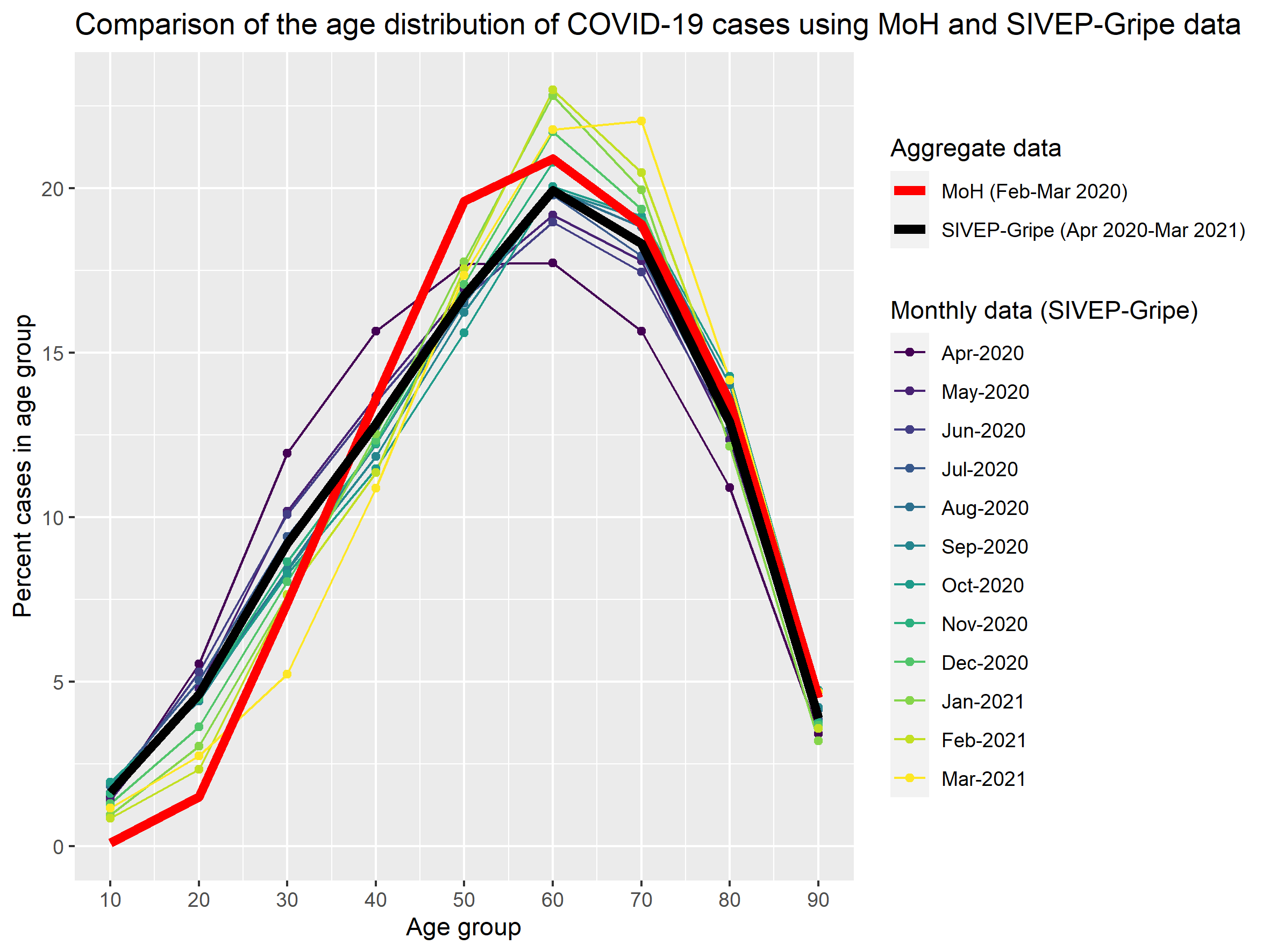
