## Supplementary material for "Tracking the emergence of disparities in the subnational spread of COVID-19 in Brazil using an online application for real-time data visualisation: a longitudinal analysis": Sensitivity analyses

**Supplementary Materials**

**Supplementary table S1 - Sensitivity analysis for time to arrival - Starting incidence = 1 case / 10000 - Epidemic arrival incidence = 5 cases / 10,0000**

| **Characteristic of the municipality** | | **Frequency** | **Median value (Interquartile range)** | **Unadjusted (univariable) model estimates**  **[95% CI]** | **Adjusted (multivariable) model estimates**  **[95% CI]** |
| --- | --- | --- | --- | --- | --- |
| **Geographic region** | **Central West** | 444 |  |  |  |
|  | **North** | 449 |  | -30·4 [-34·4,-26·5] | -30.4 [-34.4,-26.5] |
|  | **North East** | 1775 |  | -4·8 [-8·1,-1·6] | -4.8 [-8.1,-1.6] |
|  | **South** | 1157 |  | 16·2 [12·9,19·6] | 16.2 [12.9,19.6] |
|  | **South-East** | 1653 |  | 19·7 [16·3,23·1] | 19.7 [16.3,23.1] |
| **log population density (population/km^2^)** | |  | 3·20  (2·45 - 3·96) | -5·6 [-6·2,-5·0] | -7·6 [-8·2,-6·9] |
| **Percentage of residences with piped water** | |  | 72·30  (56·29 - 84·59) | -0·0 [-0·1,-0·0] | 0·0 [-0·1,0·0] |
| **Percentage of residences with piped sewage or septic tanks** | |  | 37·70  (12·75 - 70·25) | -0·0 [-0·1,-0·0] | -0·2 [-0·2,-0·1] |
| **log travel time by road to most populous municipality in the state( hours)** | |  | 2·96  (2·34 - 3·44) | -1·0 [-2·3,0·3] | 2·4 [1·2,3·7] |
| **Social Development Index** | |  | 0·25  (0·22 - 0·27) | -113·8 [-134·0,-93·5] | -91·8 [-110·6,-72·9] |

**Supplementary table S2 - Sensitivity analysis for time to arrival - Starting incidence = 1 case / 10000 - Epidemic arrival incidence = 15 cases / 10,0000**

| **Characteristic of the municipality** | | **Frequency** | **Median value (Interquartile range)** | **Unadjusted (univariable) model estimates**  **[95% CI]** | **Adjusted (multivariable) model estimates**  **[95% CI]** |
| --- | --- | --- | --- | --- | --- |
| **Geographic region** | **Central West** | 444 |  |  |  |
|  | **North** | 449 |  | -34·3 [-39·4,-29·3] | -36.7 [-41.8,-31.7] |
|  | **North East** | 1775 |  | -15·1 [-19·1,-11·1] | -2.7 [-6.8,1.5] |
|  | **South** | 1157 |  | 6·8 [2·6,11·0] | 24.8 [20.6,29.1] |
|  | **South-East** | 1653 |  | 7·5 [3·4,11·5] | 31.8 [27.5,36.1] |
| **log population density (population/km^2^)** | |  | 3·20 (2·45 - 3·96) | -5.9 [-6.6,-5.1] | -8·9 [-9·7,-8·1] |
| **Percentage of residences with piped water** | |  | 72·30 (56·29 - 84·59) | -0.0 [-0.1,0.0] | 0·0 [-0·1,0·1] |
| **Percentage of residences with piped sewage or septic tanks** | |  | 37·70 (12·75 - 70·25) | 0.0 [-0.0,0.0] | -0·2 [-0·2,-0·1] |
| **log travel time by road to most populous municipality in the state( hours)** | |  | 2·96 (2·34 - 3·44) | -0.9 [-2.6,0.8] | 4·4 [2·7,6·0] |
| **Social Development Index** | |  | 0·25 (0·22 - 0·27) | -153.0 [-178.8,-127.2] | -120·0 [-144·1,-95·9] |

**Supplementary table S3 - Comparison of the point estimates in the Tobit regression sensitivity analysis for covariates associated with the outcome (time to epidemic arrival)**

| **Covariate** | | **Point estimate of the regression effect estimate for different definitions of the threshold incidence for epidemic arrival (cases/10000)** | | | **Difference between thresholds of 10 and 5 cases per 10000**  **(% change)** | **Difference between thresholds of 10 and 15 cases per 10000**  **(% change)** |
| --- | --- | --- | --- | --- | --- | --- |
|  |  | **5** | **10** | **15** |  |  |
| **Geographic region** | **North** | -30·4 | -34·0 | -36·7 | -3·6(10·6) | 2.7(-7.9) |
|  | **North East** | -4·8 | -3·6 | -2·7 | 1·2(-33·3) | -0.9(25.0) |
|  | **South** | 16·2 | 20·8 | 24·8 | 4·6(22·1) | -4.0(-19.2) |
|  | **South-East** | 19·7 | 26·9 | 31·8 | 7·2(26·8) | -4.9(-18.2) |
| **log population density (population/km^2^)** | | -7.6 | -8·5 | -8·9 | -0·9(10·6) | 0·4(-4·7) |
| **Percentage of residences with piped sewage or septic tanks** | | 0·0 | 0·0 | 0·0 | 0·0(0·0) | 0·0(0·0) |
| **log travel time by road to most populous municipality in the state( hours)** | | -0·2 | -0·2 | -0·2 | 0·0(0·0) | 0·0(0·0) |
| **Social Development Index** | | 2·4 | 3·6 | 4·4 | 1·2(33·3) | -0·8(-22·2) |
| **log population density (population/km^2^)** | | -91·8 | -111·1 | -120·0 | -19·3(17·4) | 8·9(-8·0) |

**Supplementary table S4 - Sensitivity analysis for Rt mean estimation Summary of unadjusted and adjusted multivariate linear regression model for the association of Mean Rt (Rt mean calculated over a window from 30 to 100 days after 10 cases were reported in the municipality)**

^1^ As model contains an interaction between geographic region and start day , these estimates represent the effect size in the reference strata of the over variable

| **Characteristic of the municipality** | | **Median value (Interquartile range)** | **Freq** | **Unadjusted (univariable) model estimates**  **(95% CI) ^2^** | **Adjusted (multivariable) model estimates**  **(95% CI) ^2^** |
| --- | --- | --- | --- | --- | --- |
| **Geographic Region** | **Central West** |  | 225 | 1 | - ^1^ |
|  | **North** |  | 314 | 1·373 [1·203,1·565] | - ^1^ |
|  | **North East** |  | 992 | 1·274 [1·140,1·423] | - ^1^ |
|  | **South** |  | 462 | 1·134 [1·003,1·283] | - ^1^ |
|  | **South-East** |  | 743 | 1·246 [1·112,1·398] | - ^1^ |
| **Date of local epidemic start**  **(standardised incidence > 1 case per 10000)** | **14-Mar to 1-May**  **2020** |  | 868 | 1 | - ^1^ |
|  | **02-May to 21-May**  **2020** |  | 907 | 0·8905 [0·830,0·955] | - ^1^ |
|  | **22-May to 6-Nov**  **2020** |  | 961 | 0·6389 [0·597,0·685] | - ^1^ |
| **Population density (log_n_) (population/km^2^)** | | 3·32 (2·52 - 4·22) |  | 1·088 [1·068,1·106] | 1·044 [1·019,1·069] |
| **Percentage of residences with piped water** | | 72·77 (56·12 - 85·21) |  | 1·004 [1·002,1·005] | 1·003 [1·001,1·005] |
| **Percentage of residences with piped sewage or septic tanks** | | 36·60 (12·45 - 70·73) |  | 1·003 [1·002,1·004] | 1·004 [1·002,1·005] |
| **Travel time (log_n_) by road to most populous municipality in the state (hours)** | | 2·96 (2·32 - 3·46) |  | 1·005 [0·964,1·049] | 0·991 [0·945,1·04] |
| **Social Development Index (SDI)** | | 0·25 (0·22 - 0·28) |  | 3·494 [1·774,6·883] | 3·102 [1·530,6·284] |

**Supplementary table S5 - Sensitivity analysis for Rt mean estimation Summary of unadjusted and adjusted multivariate linear regression model for the association of Mean Rt (Rt mean calculated over a window from 30 to 180 days after 10 cases were reported in the municipality)**

| **Characteristic of the municipality** | | **Median value (Interquartile range)** | **Freq** | **Unadjusted (univariable) model estimates**  **(95% CI) ^2^** | **Adjusted (multivariable) model estimates**  **(95% CI) ^2^** |
| --- | --- | --- | --- | --- | --- |
| **Geographic Region** | **Central West** |  | 228 | 1 | - ^1^ |
|  | **North** |  | 314 | 1·401 [1·241,1·581] | - ^1^ |
|  | **North East** |  | 993 | 1·266 [1·143,1·401] | - ^1^ |
|  | **South** |  | 471 | 1·355 [1·212,1·516] | - ^1^ |
|  | **South-East** |  | 754 | 1·326 [1·195,1·473] | - ^1^ |
| **Date of local epidemic start**  **(standardised incidence > 1 case per 10000)** | **14-Mar to 1-May**  **2020** |  | 875 | 1 | - ^1^ |
|  | **02-May to 21-May**  **2020** |  | 914 | 0·860 [0·806,0·918] | - ^1^ |
|  | **22-May to 6-Nov**  **2020** |  | 971 | 0·739[0·693,0·787] | - ^1^ |
| **Population density (log_n_) (population/km^2^)** | | 3·31 (2·52 - 4·21) |  | 1·091 [1·074,1·110] | 1·07 [1·046,1·094] |
| **Percentage of residences with piped water** | | 72·76 (56·12 - 85·18) |  | 1·004 [1·003,1·005] | 1·003 [1·001,1·004] |
| **Percentage of residences with piped sewage or septic tanks** | | 36·75 (12·45 - 70·73) |  | 1·003 [1·002,1·004] | 1·003 [1·002,1·004] |
| **Travel time (log_n_) by road to most populous municipality in the state (hours)** | | 2·96 (2·32 - 3·46) |  | 1·022 [0·984,1·063] | 1·027 [0·982,1·074] |
| **Social Development Index (SDI)** | | 0·25 (0·22 - 0·28) |  | 2·956 [1·587,5·512] | 2·651 [1·38,5·094] |

^1^ As model contains an interaction between geographic region and start day , these estimates represent the effect size in the reference strata of the over variable
